# Multi-PGS enhances polygenic prediction: weighting 937 polygenic scores

**DOI:** 10.1101/2022.09.14.22279940

**Authors:** Clara Albiñana, Zhihong Zhu, Andrew J. Schork, Andrés Ingason, Hugues Aschard, Isabell Brikell, Cynthia M. Bulik, Liselotte V. Petersen, Esben Agerbo, Jakob Grove, Merete Nordentoft, David M. Hougaard, Thomas Werge, Anders D. Børglum, Preben Bo Mortensen, John J. McGrath, Benjamin M. Neale, Florian Privé, Bjarni J. Vilhjálmsson

## Abstract

The predictive performance of polygenic scores (PGS) is largely dependent on the number of samples available to train the PGS. Increasing the sample size for a specific phenotype is expensive and takes time, but this sample size can be effectively increased by using genetically correlated phenotypes. We propose a framework to generate multi-PGS from thousands of publicly available genome-wide association studies (GWAS) with no need to individually select the most relevant ones. In this study, the multi-PGS framework increased prediction accuracy over single PGS for all included psychiatric disorders and other available outcomes, with prediction R2 increases of up to 9-fold for attention-deficit/hyperactivity disorder (ADHD) compared to a single PGS. We also generate multi-PGS for phenotypes without an existing GWAS and for case-case predictions, with up to 15-fold increases in prediction accuracy. We benchmark the multi-PGS framework against other methods and highlight its potential application to new emerging biobanks.

## Main

Although polygenic scores (PGS) have high potential for clinical use^1–3^, they are currently underpowered for many applications regarding disease prediction and risk stratification. The predictive performance of PGS is largely determined by four factors: the sample size of the GWAS used for training the score, the proportion of causal variants and the heritability of the phenotype, as well as heterogeneity between GWAS and target samples, including differences in genetic ancestry^4–6^. Increasing the number of samples for a phenotype is costly and takes time, but a possible alternative is to use genetically correlated phenotypes to increase the effective sample size at no cost^1,7–14^. Most of the multi-phenotype approaches require an available GWAS for the disease of interest and to pre-define a set of relevant GWAS summary statistics, based on prior information about the disease. Our proposed multi-PGS strategy works around those two conditions. This multi-PGS is trained on thousands of different PGS such as for health outcomes, body measurements, and behavioral phenotypes which are not necessarily genetically correlated with the outcome. The multiple PGS are combined using penalized regression models^7,11^, which selects the specific PGS that increase prediction accuracy of the disease of interest.

Previously proposed multi-PGS have either required individual-level genotype and phenotype validation data for each PGS in the model^7,11^, or the inclusion of multiple PGS for the same GWAS summary statistics file corresponding to different p-value thresholds or proportions of causal variants^15^ (i.e. the PGS model hyper-parameters). For the latter, the number of PGS included in the multi-PGS prediction can become very large (e.g. 10 for each GWAS summary statistic number), which increases the number of validation samples required to fit the multi-PGS model, and limits the number of PGS that can practically be included in the final multi-PGS.

Recent advances in PGS methods allow us to generate PGS for a phenotype without requiring validation data to tune the hyper-parameters^16–20^. This development has two major implications in the context of multi-PGS. First, individual-level genotype validation data for each of the correlated phenotypes included in the multi-PGS is no longer necessary because selecting the best-performing hyper-parameters is no longer needed. Second, PGS for any genetically correlated phenotype, even those not available in the target data, can now be included more easily in the multi-PGS, significantly expanding the set of phenotypes one can study. Therefore only one PGS per phenotype needs to be included in the multi-PGS, then the only practical limitation is effectively the number of individual-level samples available for the desired phenotype, which is constantly growing for biobank data.

In this study, we propose and evaluate a specific framework that leverages these key PGS developments to construct more powerful and generalisable multi-PGS. We construct the multi-PGS in two steps. First, we construct a broad PGS library for hundreds of diverse phenotypes from publicly available GWAS summary statistics (without the need for PGS specific validation data). Second, we use this PGS library to fit a prediction model for any of the phenotypes available in a biobank. As a prediction model for combining the PGS, we compare either using both L1 penalized regression and scalable gradient boosted trees (XGBoost)^21^. XGBoost has the benefit or advantage that it can capture interactions in the model (e.g., interactions between covariates, between PGS, as well as between PGS and covariates).

We apply our multi-PGS framework to the Lundbeck Foundation Initiative for Integrative Psychiatric Research (iPSYCH)^22,23^, one of the largest datasets on the genetics of major psychiatric disorders. These disorders are genetically correlated with many other psychiatric and neurological disorders as well as other behavioral phenotypes^24,25^, which are precisely the circumstances under which the proposed multi-PGS might boost the polygenic prediction accuracy. We benchmark the multi-PGS against each phenotype’s respective single PGS prediction and compare it with an existing re-weighting method that does not train on individual-level data, wMT-SBLUP^8^.

Although the iPSYCH cohort has been designed around psychiatric disorders, the study individuals can be linked to the National Danish Registers^22,23^, making it possible to generate multi-PGS for any phenotype captured in these registers. We demonstrate that multi-PGS improves prediction accuracy results for a range of different diseases, subtypes and phenotypes for which no GWAS summary statistics currently exist (e.g., birth measurements and case-case classification). Our goal is to showcase the approach to develop multi-PGS and its potential advantage to be applied to new emerging biobank data.

## Results

### Overview of method

Here we outline the framework used for generating the proposed multi-PGS. Any GWAS summary statistics for any phenotype can be used to generate a PGS to be included in the multi-PGS, even if no validation data are available for these. This is made possible by using LDpred2-auto^16^, which does not require a validation set. First, a library of GWAS summary statistics is generated, making sure it has the same genetic ancestry and the samples used do not overlap with the target data. Then, a PGS is generated for each GWAS summary statistic, where the PGS are all standardized (i.e., mean-centered with variance 1). The PGS library construction is described in detail in the methods section and supplementary text. To combine the PGS in the library into a multi-PGS, the target individual-level genotypes together with the phenotype of interest and covariates are required (here sex, age and 20 first PCs). The prediction accuracy of the multi-PGS is evaluated through a cross-validation scheme and it is benchmarked to the prediction accuracy of single PGS and different PGS re-weighting methods^8^. As we will show, this enables us to train enhanced *polygenic scores*, even for phenotypes for which there is no external GWAS available. The overview of the framework and the analysis strategy is depicted in **Figure 1**.

**Figure 1:**
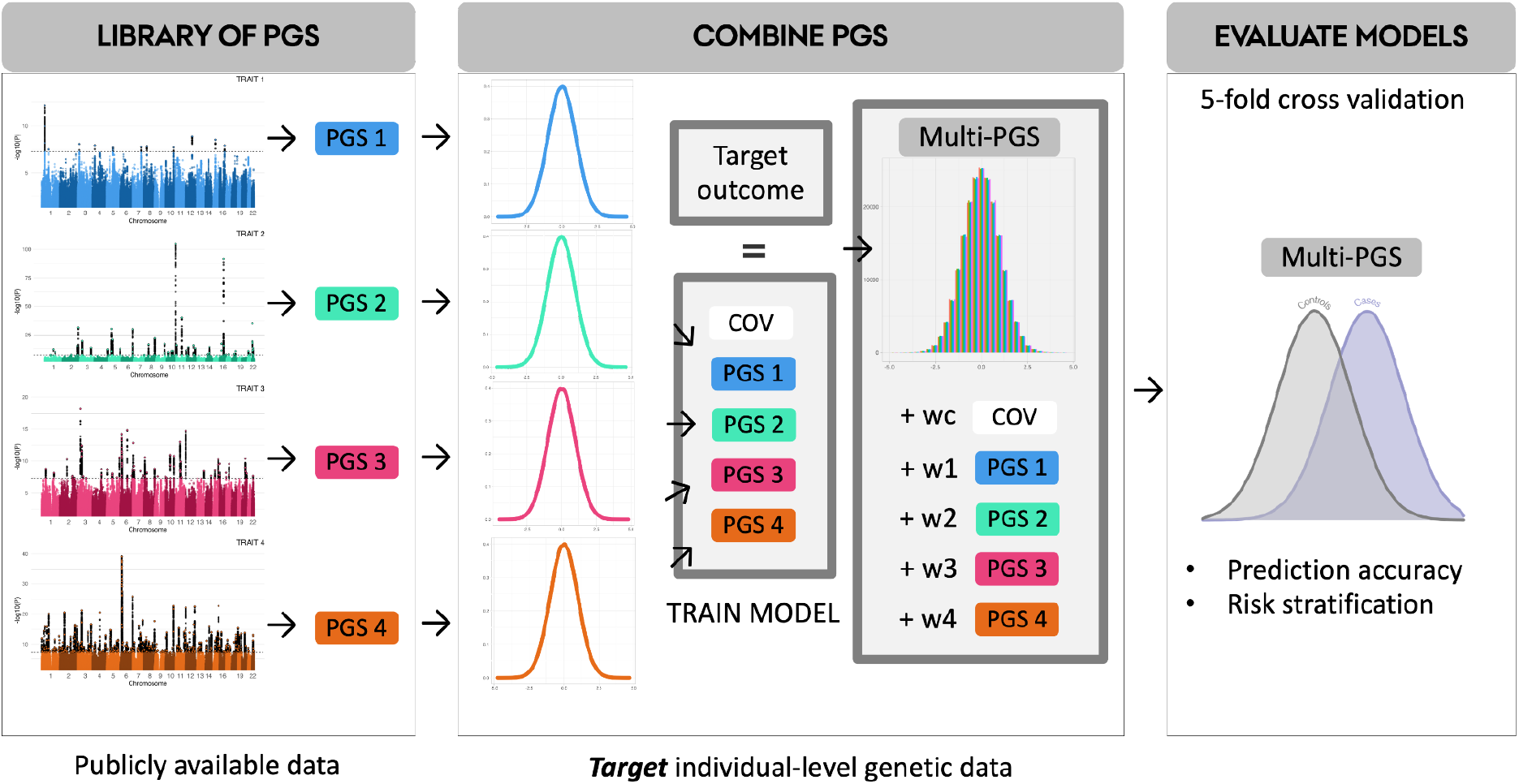
Overview of the multi-PGS framework.

### Linear and non-linear combinations of PGS give comparable prediction results

We first studied the relationship between the covariates (sex, age and first 20 PCs) and the 937 PGS by comparing the performance of linear models (lasso penalized regression; multiPGS_lasso) and non-linear models (boosted gradient trees: multiPGS_XGBoost) to predict the following 6 major psychiatric disorders: attention-deficit/hyperactivity disorder (ADHD), affective disorder (AFF), anorexia nervosa (AN), autism spectrum disorder (ASD), bipolar disorder (BD) and schizophrenia (SCZ). We used a model including a single PGS for the largest available GWAS for each psychiatric disorder as the standard reference (ST3). In terms of variance explained, the multi-PGS models increased the mean R2 4-fold on average over the single GWAS PGS for all disorders, with up to 9-fold improvement for ADHD and ASD (Figure 2A), from 0.8 to 7.5 and from 0.2 to 2, respectively. Compared to the middle risk score quintiles, multi-PGS generally increased the log odds ratio over the single GWAS PGS (Figure 2B).

**Figure 2.**
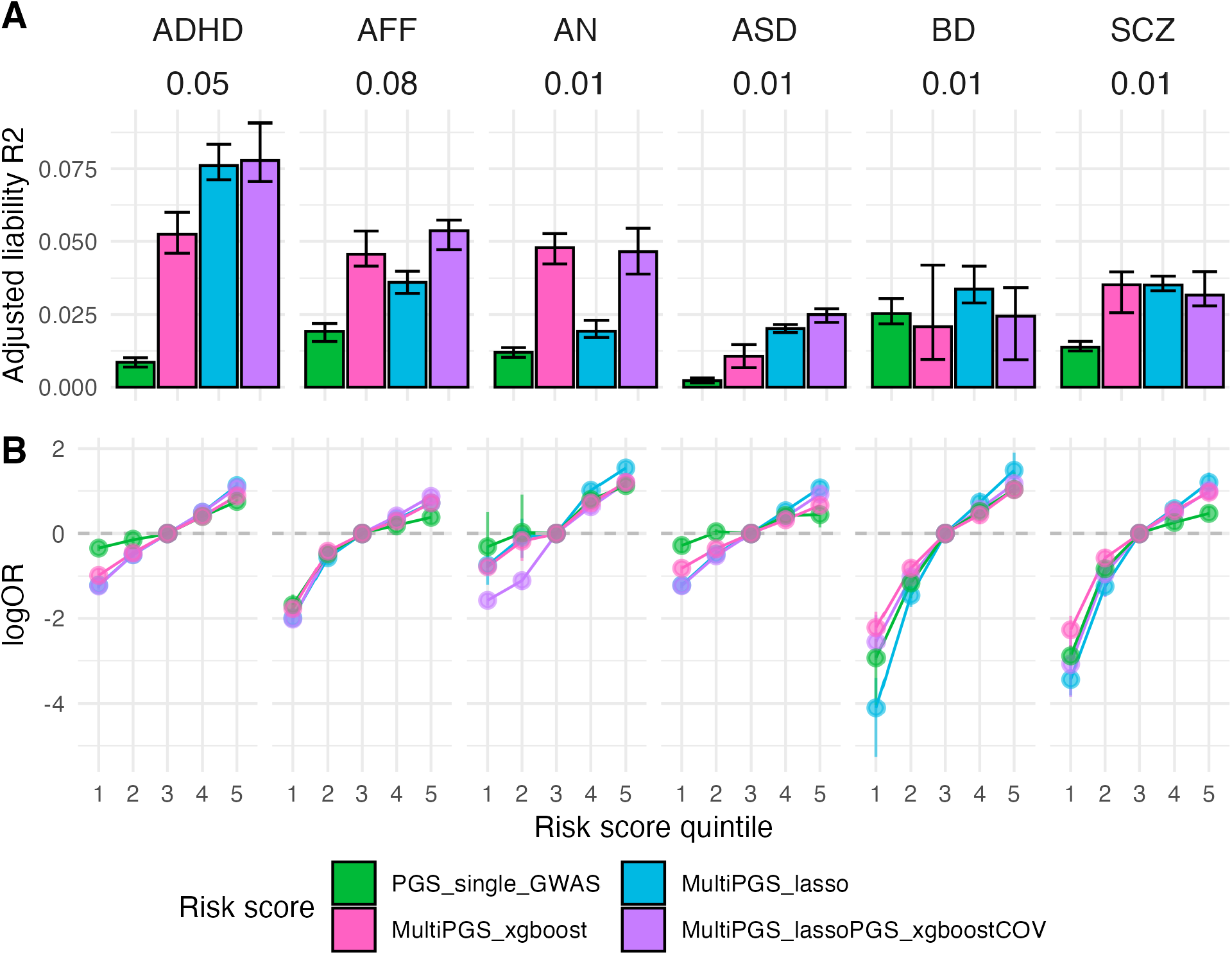
Performance of the different risk score models including covariates. Comparison between the per-disorder attention-deficit/hyperactivity disorder (ADHD), affective disorder (AFF), anorexia nervosa (AN), autism spectrum disorder (ASD), bipolar disorder (BD) and schizophrenia (SCZ) single GWAS PGS (specific details on ST3) and the multi-PRS models trained with 937 PGS in terms of A) liability adjusted R2 and B) log odds ratios compared to the middle risk score quintiles. All models included sex, age and first 20 PCs for training the different PGS weights and calculating the risk score on the test set in a 5-fold cross-validation scheme. 95% confidence intervals were calculated from 10,000 bootstrap samples of the mean adjusted R2 or logOR, where the adjusted R2 was the variance explained by the full model after accounting for the variance explained by a logistic regression covariates-only model as R2_adjusted = (R2_full - R2_cov) / (1 - R2_cov). Prevalences used for the liability are shown beneath each disorder label and case-control ratios are available on ST3.

In terms of which multi-PGS model performed better at combining the variables, the results appear to be disorder specific. For ADHD, ASD and BD the lasso multi-PGS increased the mean prediction R2 over the XGBoost multi-PGS. On the contrary, for AFF and AN the XGBoost multi-PGS increased the mean prediction R2 over lasso multi-PGS. For SCZ, there was no difference in variance explained by the two models.

We further investigated if the increase in prediction of the XGBoost multi-PGS over lasso multi-PGS was driven by the nonlinear combination of covariates alone, and if it was independent from the PGS combination. In practice, we obtained an XGBoost risk score for the covariates and fitted it as an additional variable in the lasso model, together with the 937 PGS. The mean variance explained by this mixed multi-PGS was comparable to the lasso multi-PGS for ADHD, ASD, BD and SCZ, while it was comparable to the mean variance explained by the XGBoost multi-PGS for AFF and AN. This demonstrated that the non-linear combination was only beneficial at the covariate level and not at the PGS-level.

Less pronounced differences were observed in terms of the mean area under the curve (AUC) prediction (SF5), indicating that both models are similar in terms of classification. In terms of quintile odds ratio, the lasso multi-PGS was generally the best at separating the top 20% to the middle quintile, even for the models where the maximum mean variance was explained by the XGBoost multi-PGS (Figure 2B). Since the two multi-PGS models provided relatively similar results, we continued further analyses on the psychiatric disorders considering only the lasso multi-PGS, as weights from linear models are more interpretable.

### Comparison between single PGS and multi-PGS predictors

Next, we investigated which PGS in the multi-PGS model were the ones contributing the most to increasing prediction accuracy. The number of non-zero PGS in each multi-PGS model ranged from 10 to 154, where the number of PGS included correlated with the number of samples in the training set. However, weights were generally very small, with few PGS with an absolute lasso weight larger than 0.01 (SF6-SF11). We compared the prediction accuracy of a multi-PGS with all non-zero weights to a simplified lasso multi-PGS that included only PGS with an absolute lasso weight larger than 0.01, multi-PGS_lasso_s (Figure 3, number of included PGS in the figure), showing very similar mean R2. Prediction estimates for the top 5 PGS in each model ranked by their lasso weights are also shown in Figure 3, with the top PGS generally contributing to half of the prediction accuracy of the multi-PGS models.

**Figure 3.**
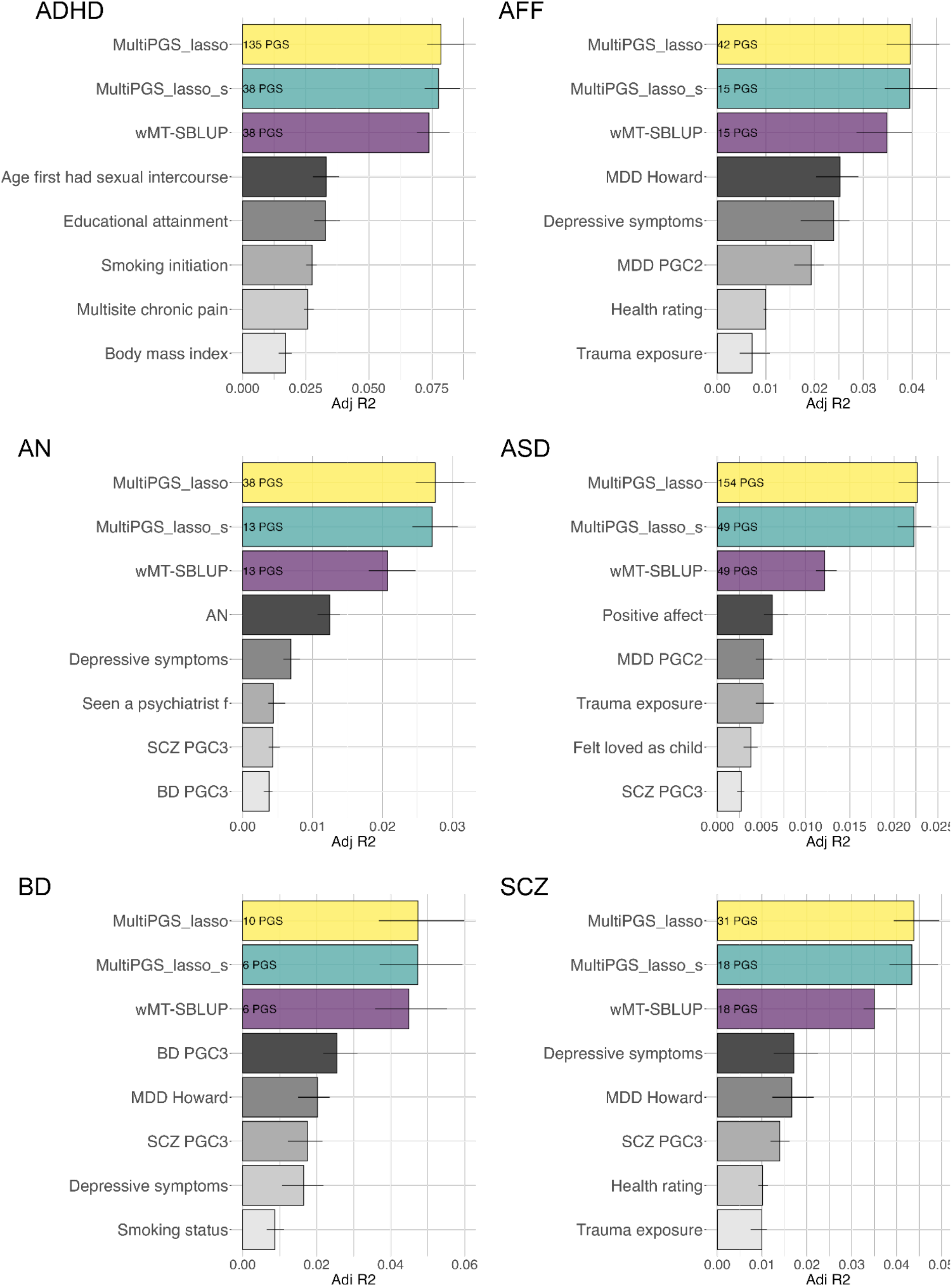
Comparison between single-phenotype and multi-phenotype PGS (multi-PGS and wMT-SBLUP). Mean liability adjusted R2 estimates between attention-deficit/hyperactivity disorder (ADHD), affective disorder (AFF), anorexia nervosa (AN), autism spectrum disorder (ASD), bipolar disorder (BD) and schizophrenia (SCZ) and multi-phenotype predictors (colored bars, multiPGS_lasso, multiPGS_lasso_s, wMT-SBLUP) or single-phenotype PGS (grayscale bars, single LDpred2-auto PGS). The adjusted R2 estimates are the mean of the 5-fold cross-validation training-testing subsets. CI were calculated from 10k bootstrap samples of the mean. The numbers inside each multi-phenotype predictor correspond to the number PGS included in each model. Both the simplified multi-PGS (multiPGS_lasso_s) and wMT-SBLUP predictors were calculated by keeping the top PGS with an absolute lasso weights > 0.01 from the full multi-PGS, including the top 5 shown in the figure.

By using the lasso regression as a feature selection algorithm (only selecting the ones with large weights), it was feasible to compare the prediction accuracy to another multi-PGS model, wMT-SBLUP^8^, as this method required to calculate a genetic correlation matrix between all PGS in the model. The PGS weights in wMT-SBLUP are calculated as a function of the GWAS summary statistics’ sample size, SNP-heritability and genetic correlation (SF12 contains an overview of these parameters for the top 10 PGS in each model). The simplified multi-PGS showed consistently higher mean R2 than wMT-SBLUP, even though they both contained the same number of PGS.

### Re-weighting hundreds of external PGS increases prediction over training only on the individual-level data

We then compared the difference in the prediction models of using only one type of data (individual-level or GWAS summary statistics) vs. re-weighting the set of PGS, which uses both types of data. First, we benchmarked the lasso multi-PGS against the single PGS trained on the external largest available GWAS summary statistics and the single PGS trained on the individual-level genotype and phenotype information (BLUP PGS, details in methods section). As shown previously^26^, the BLUP PGS vs. single GWAS PGS varied in the relative proportion of variance explained according to the psychiatric disorder, as they are largely dependent on the training sample sizes and genetic correlation (Figure 4A). We observed these large differences also in terms of log OR of separating the top 10% to the bottom 10% of the sample (Figure 4B).

**Figure 4.**
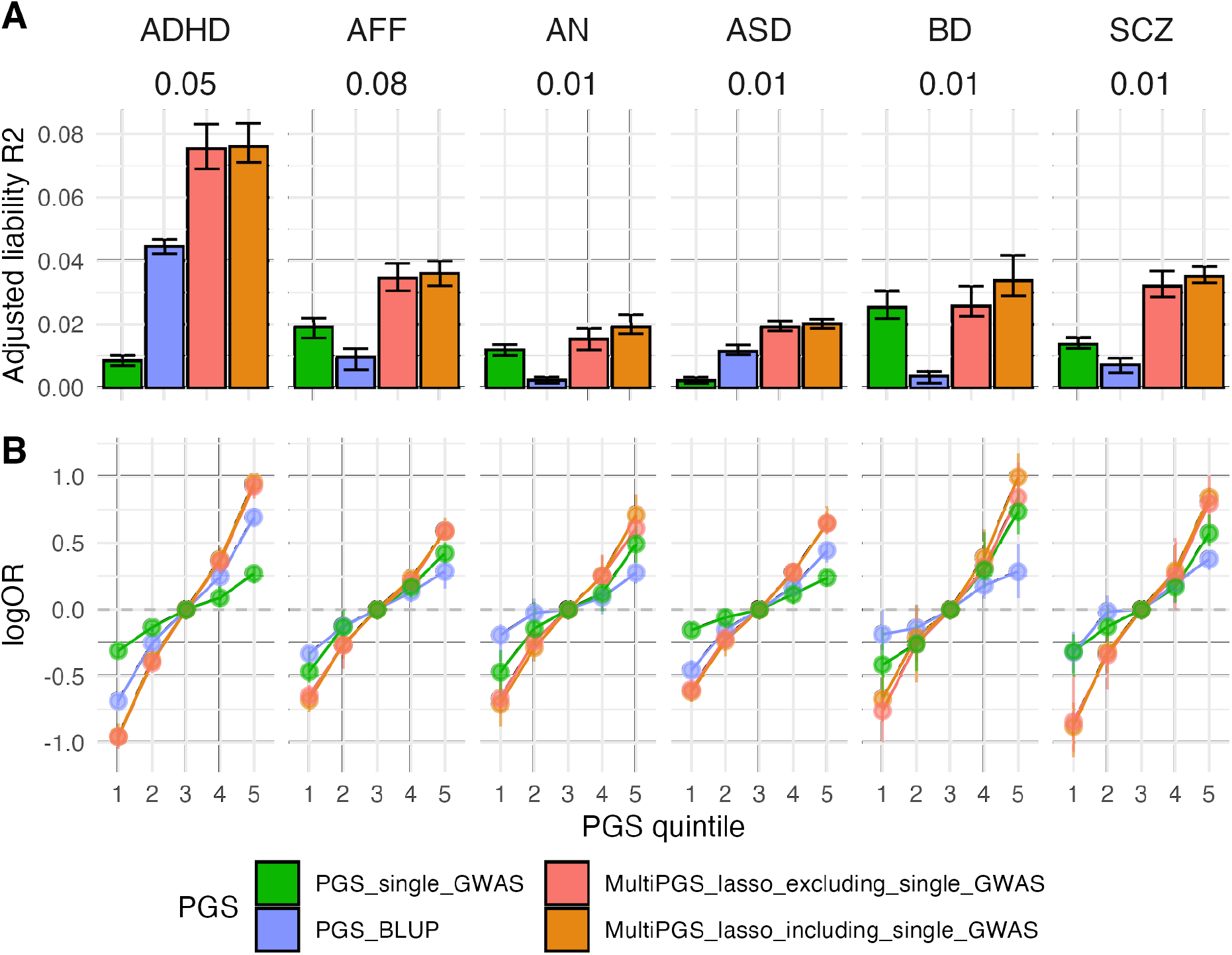
Performance of the different PGS models trained with different data. Comparison between the per-disorder attention-deficit/hyperactivity disorder (ADHD), affective disorder (AFF), anorexia nervosa (AN), autism spectrum disorder (ASD), bipolar disorder (BD) and schizophrenia (SCZ) single GWAS PGS (PGS_single_GWAS) (Details on ST3), the per-disorder BLUP PGS and the multi-PGS models in terms of A) liability adjusted R2 and B) log odd ratios. The multiPGS_lasso_excluding_single_GWAS represents the PGS where the specific single GWAS PGS was removed from the set of 937 PGS. All models were adjusted for sex, age and first 20 PCs. The adjusted liability R2 shows the mean of the 5-fold cross-validation training-testing subsets. CI were calculated from 10k bootstrap samples of the mean adjusted R2 or logOR, where the adjusted R2 was the variance explained by the full model after accounting for the variance explained by a logistic-regression covariates-only model as R2_adjusted = (R2_full - R2_cov) / (1 - R2_cov). Prevalences used for the liability are shown beneath each disorder label and case-control ratios are available on ST3.

Although we observed a clear prediction accuracy increase of multi-PGS over single PGS, there are not always available external GWAS summary statistics for the specific phenotype to generate single PGS. To test the influence in the multi-PGS prediction accuracy of having/not having a PGS for the desired phenotype in the PGS library, we compared two types of lasso multi-PGS: one trained on the set of 937 PGS and one trained using the set of 937 minus the specific PGC external PGS for each psychiatric disorder. The difference in mean prediction R2 was not significant between both multi-PRS for any psychiatric disorders (Figure 4A) and the multi-PGS that did not contain the disorder’s external PGS was not worse at separating the risk deciles in terms of log OR (Figure 4B).

### Generating multi-PGS from register-based phenotypes

Finally, we extended the multi-PGS results to other phenotypes defined in the danish national registers in the overlapping samples with iPSYCH to showcase its potential to generate polygenic scores in a biobank framework. First, we selected 62 ICD10 codes from the Danish Psychiatric Central Research Register^27^ with at least 500 diagnosed cases and compared the prediction performance of the multi-PGS lasso to the multi-PGS XGBoost. Similarly to the results for the main 6 psychiatric disorders, the mean prediction R2 for both multi-PGS was on average the same, but ultimately depended on the disorder (SF13). We emphasize that both these phenotypes all nested within the psychiatric disorder cases and population cohort of iPSYCH case-cohort design (ST4).

To expand the comparison to external data, we selected 15 phenotypes from four different categories; a) other ICD10 codes with available GWAS summary statistics, b) other ICD10 codes with not known available GWAS summary statistics / disorder sub-phenotypes, c) continuous phenotypes from the Medical Birth Register^28^ and d) case-case predictions. For the last category, we explored two example pairs of disorders with a high degree of comorbidity. First, we excluded the cases with both disorders and re-codified each single disorder case as 0 or 1. We then paired each phenotype with an appropriate PGS from the PGS library (details on ST5) and compared the mean R2 prediction performance of the multi-PGS lasso and single GWAS PGS. The results varied greatly for each phenotype, but there were large increases in prediction accuracy of the multi-PGS for some phenotypes (Figure 5). For example, we show 15-fold for ADHD/ASD, 9-fold for Asperger’s syndrome, 6-fold for Substance abuse disorders, but null R2 for other phenotypes (APGAR score, gestational age, BD/MDD. The multi-PGS for the disorder pair ASD/ADHD showed the largest prediction R2 of all examined multi-PGS, explaining 12% of the variation.

**Figure 5.**
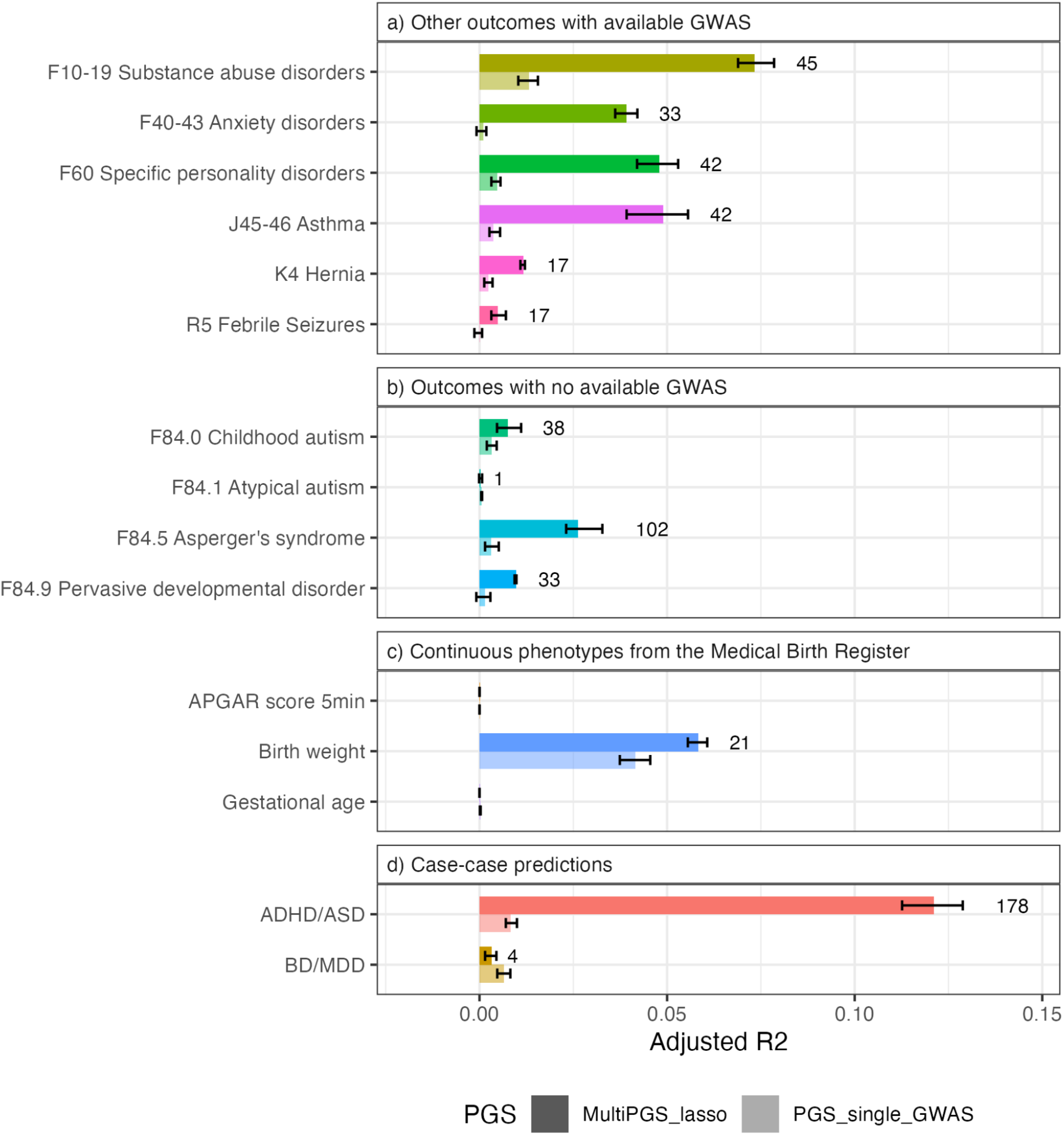
Examples of the prediction accuracy of multi-PGS on register-based phenotypes. Comparison between a per-phenotype single GWAS PGS (for the phenotypes with available external GWAS summary statistics or the most similar PGS in the library, details on ST5) and the models trained with 937 PGS in terms of adjusted R2. All models included sex, age and first 20 PCs for training the different PGS weights and calculating the risk score on the test set in a 5-fold cross-validation scheme. CI were calculated from 10,000 bootstrap samples of the mean adjusted R2, where the adjusted R2 was the variance explained by the full model after accounting for the variance explained by a logistic regression covariates-only model as R2_adjusted = (R2_full - R2_cov) / (1 - R2_cov). The number next to the multiPGS bar indicates the number of non-zero lasso mean weights for the 5 cross-validation subsets.

## Discussion

Here we have proposed a multi-PGS framework derived from nearly one thousand GWAS summary statistics for different phenotypes, and showed that it increased the accuracy of PGS for psychiatric disorders. Multi-PGS explained a larger proportion of the SNP-heritability (a four-fold increase on average for the main psychiatric disorders over a PGS trained on the target outcome only) and stratified the population in more distinct risk groups than single-phenotype PGS. The disorder with the largest increase in prediction accuracy from multi-PGS was ADHD, with a 9-fold increased prediction R2 over the single PGS (corresponding to ∼40% of the SNP h2^29^). This is mostly due to the inclusion of various behavioral PGS in our library (educational attainment, smoking status, etc.) with very large sample sizes and large genetic correlation with ADHD.

While the compared *linear* multi-PGS (lasso) and *non-linear* multi-PGS (XGBoost) resulted in similar prediction accuracies, the improved performance of the non-linear models for affective disorder and anorexia nervosa was due to the presence of non-linear covariate interactions between sex and age, as previously reported^30^. We therefore focused the analyses on the lasso multi-PGS, as those results have easier interpretability.

We benchmarked our multi-PGS prediction results against a PGS trained on external GWAS summary statistics for each of the six major psychiatric disorders respectively. The multi-PGS always resulted in a more accurate prediction, even when the phenotype itself was not included in the PGS library. When benchmarked against wMT-SBLUP^8^, another PGS method trained on GWAS summary statistics for multiple phenotypes, the multi-PGS also resulted in a more accurate prediction. The improvement in prediction accuracy is likely due to the fact that while wMT-SBLUP bases its weights on the genetic correlation to the external GWAS summary statistics for each phenotype, multi-PGS is trained directly on the individual-level samples, allowing multi-PGS to better tailor the weights to the cohort. Interestingly, the multi-PGS still obtained much more accurate predictions by re-weighting hundreds of external PGS. This result suggests that as both the number of genome-wide association studies and their sample size grows, training multi-PGS will become more feasible.

Finally we showcased how multi-PGS can be used to train predictors for any phenotype of interest, as long as one has *sufficient* individual-level genetic data available with the phenotype of interest. The multiPGS predictors do not require PGS for the target phenotype of interest to be available in the PGS library used. We demonstrated in practice how these multi-PGS could be generated for various psychiatric sub-diagnoses and case-case prediction, including classification of ADHD and ASD with relatively high prediction accuracy.

This study and the multi-PGS approach has several limitations. First, as we performed a 5-fold cross-validation in the iPSYCH (individual-level) data when training and testing the multi-PGS, it is possible that some of the prediction accuracy gain is due to overfitting. However, the multi-PGS also resulted in multiple null predictions despite having large training sample sizes (APGAR score and gestational age) suggesting that overfitting is small. To further avoid overfitting we restricted the analyses to a set of unrelated individuals of European ancestry to control for population structure. Second, controlling for sample overlap between external and internal data, which can lead to overfitting^31^, becomes both difficult and important when considering thousands of GWAS summary statistics. We addressed this manually by checking the GWAS summary statistics, but an automatization of this step (e.g. using bivariate LDSC^32^) could help streamline this procedure. Third, the resulting multi-PGS is potentially predicting a subset of individuals in the case group enriched or comorbid with a phenotype with large genetic correlation to the PGS phenotypes, but not the disorder itself. Therefore, although multi-PGS can improve prediction accuracy, it should not be used to estimate genetic correlations or study genetic overlap. Fourth, we have not explored whether including a PRS trained on individual-level data could improve the prediction further, as suggested by a previous study^26^. Fifth, we have not explored how generalisable the multi-PGS are across different ancestry groups, but we expect the R2 prediction accuracy to decay with genetic distance between training and testing, as previously shown for PGS^6,33^. However, as more individuals of non-European ancestry are included in GWAS, the multi-PGS based on the resulting summary statistics may be able to improve cross-ancestry prediction.

The same multi-PGS framework could be applied to other types of biological data summary statistics, like GWAS for brain or cardiac images^34,35^, gene expression^36^, and/or protein levels^37^. Combining different types of data into a multi-PGS could potentially improve or quantify the importance for prediction of the different data types. The framework can also make use of published PGS variant weights (e.g. from the PGS catalog^38^) instead of deriving the PGS from the GWAS summary statistics.

In this study, by leveraging nearly a thousand external PGS, we show how we can increase the polygenic prediction further without the need to genotype more individuals of a specific phenotype. We think this multi-PGS framework has a lot of potential for new emerging biobanks or register-based genetic cohorts to generate PGS for every available phecode or defined phenotype in their system, both because of the ever growing set of publicly available GWAS summary statistics and, as they are new, these biobanks do not have the issue of sample overlap with the external GWAS summary statistics used.

## Online methods

### PGS library construction

A detailed description of the PGS library construction and file filtering process is provided as Supplementary Text and all code used will be available. A number of resources were used to obtain an initial list of GWAS summary statistics for generating the PGS library. The majority of the GWAS summary statistics were downloaded from publicly available databases (GWAS Catalog^39^, GWAS Atlas^40^, the Psychiatric Genomics Consortium Website (https://www.med.unc.edu/pgc). For the specific PGC GWAS summary statistics where iPSYCH was used in the discovery dataset, we used in-house GWAS results where these samples were excluded from the calculation. The GWAS files were specifically selected to be based on European ancestry individuals, to not be overly redundant (only the latest GWAS for each same phenotype) and to not contain iPSYCH samples. From an original list of 6,206 files (ST1), this filtering resulted in 1,377 files to download.

We developed a pipeline for downloading, parsing, reformatting and doing a quality control filter on the list of files. We created a GWAS summary statistic column-name library (SF1) to ease file parsing, and after re-formatting and removing corrupted files, this step resulted in 1,005 files. We restricted the number of SNPs to the overlap of the iPSYCH imputed variants with the HapMap3 variants and the LD reference provided by LDpred2, resulting in a maximum of 1,053,299 SNPs per GWAS summary statistics file. Filtering SNPs with a large discrepancy in standard deviation between the genotyped/imputed data and the GWAS summary statistics can increase the robustness and prediction accuracy of PGS^41^. For each file, we created a QC plot for visual inspection of the QC SNP filtering (Example in SF2). The set of 952 GWAS summary statistics that passed QC and kept over 200,000 SNPs were used to derive PGS with LDpred2-auto ^16^.

Polygenic scores were derived using LDpred2-auto, a method within the LDpred2 framework^16^ that does not require a validation dataset to fit the hyperparameters (SNP-h2; SNP-based heritability estimate and p; proportion of causal SNPs), but these are fitted as part of the Gibbs sampler instead. We used the provided European-ancestry independent LD blocks as reference panel^41^. For each GWAS summary statistics file, LDpred2-auto was run with 30 Gibbs sampler chains, 800 burn-in iterations and 400 iterations. The SNP-h2 initial value was set to the LD score regression estimate^42^ from the GWAS summary statistics after. Each of the chains was initialized with a different prior for the proportion of causal variants: [1e-4, 0.9] in log scale (example plot for a chain in SF3). Chains were filtered according to the recommendation in the LDpred2 tutorial, and effect sizes of chains kept were averaged.

After running LDpred2-auto on the QC’d file set and post-processing, we were left with 937 PGS. This constitutes the final PGS library and all information on its GWAS summary statistics meta-data, number of SNPs per file on each step, number of chains on the final PGS and estimates of SNP-h2 and p can be found in ST2. A plot comparing the LDSC and LDpred2-auto SNP-h2 estimates can be found at SF4.

### iPSYCH data

#### Genotypes and imputation

The iPSYCH 2015 case-cohort sample is a genotyped dataset from neonatal dried blood spots (DBS) nested within the entire Danish population born between 1981 and 2008, including 1,657,449 persons. After genotyping and sample quality control (described in detail elsewhere ^22,23^), it includes 92,765 individuals diagnosed with a major psychiatric disorder i.e. attention-deficit/hyperactivity disorder (ADHD), affective disorder (AFF), autism (ASD), schizophrenia (SCZ) and bipolar disorder (BD). We also included the anorexia nervosa (AN; ANGI-DK) samples from the Anorexia Nervosa Genetics Initiative (ANGI)^43^, as they were samples within the same framework as iPSYCH 2015. The dataset also includes 42,912 individuals randomly sampled from the same birth cohort, making it representative of the general Danish population. The genotype data was imputed using the Haplotype Reference Consortium (HRC)^44^ as the reference panel and following the RICOPILI pipeline^45^. After removing SNPs with minor allele frequency (MAF) < 0.01 and Hardy-Weinberg p-value < 10-6, we restricted to the HapMap3 variants in the LDpred2 LD reference panel, resulting in 1,053,299 SNPs.

#### Principal components & relatedness

We performed principal component analysis (PCA) following Privé *et al*. ^46^ and obtained 20 PCs. The process has already been described in Albiñana *et al*. 2021 ^26^. Using the set of 20 PCs, we defined genetically homogeneous individuals as having <4.5 log distance units to the multidimensional center of the 20 PCs (calculated using the function dist_ogk from the R package bigutilsr^46,47^). We also computed the KING-relatedness robust coefficient of the sample and excluded the second of each pair with >3rd degree relatedness. We identified a set of 108,031 unrelated genetically homogeneous individuals (Danish-European ancestry), which we used for all subsequent analyses.

#### List of phenotypes and ICD10 codes

We used phenotypes from the Danish Psychiatric Central Research Register^27^ with register data available until December 2016. Except for the category of all psychiatric disorders (ICD10, F-chapter), all categories of diagnosis are given a variable name starting with a letter and followed by four digits. The first letter is the chapter of disease in the ICD10 system and the first number is the ICD10 diagnosis. The other 3 numbers are not informative of ICD10 diagnosis. We also used 3 continuous phenotypes from the Danish Medical Birth Register^28^: apgar5 (Apgar score, 5 minutes after birth), fvagt (birth weight in grams) and gest_age (gestational age in completed weeks). All used phenotypes, sample sizes and metadata are available at ST4-ST5.

#### Multi-PGS models

We used the library of 937 PGS to train multi-phenotype predictors (multi-PGS) using two different algorithms 1) L1 penalized regression (lasso) as implemented in the R package glmnet ^48^ and 2) tree gradient boosting as implemented in the XGBoost (eXtreme Gradient Boosting) algorithm in the R package xgboost ^21^. For both models, we trained a base model using only the covariates (sex, birth year and 20 PCs) and a full model using the covariates plus the 937 standarized LDpred2-auto PGS. For the base model, we used the glm function with the option family = “logistic”. For lasso, we used the function cv.glmnet from the glmnet R package with the options alpha = 0 and family = “binomial” for binary phenotypes. The covariates were not regularized by giving them a penalty factor of 0 with the option penalty.factor, while the rest of the PGS were given a penalty factor of 1. For XGBoost, we used the xgboost function from the xgboost R package with options eta = 0.01 and nrounds = 10. We used objective = “binary:logistic” for binary phenotypes and objective = “reg:squarederror” for continuous variables.

#### BLUP PGS

We computed an internal best linear unbiased prediction (BLUP) PGS trained on the individual-level data. We obtained the per-SNP prediction betas with BOLT-LMM^49,50^ (using the flag –predBetasFile) on the set of 1,118,443 HM3 SNPs on iPSYCH. Depending on the polygenicity of the phenotype, BOLT-LMM computes a mixture-of-Gaussians prior or a single-Gaussian BOLT-LMM-inf model, equivalent to best linear unbiased prediction (BLUP). In the case of psychiatric disorders, our results show that BOLT-LMM-inf is always the model selected and therefore we refer to the BOLT-LMM PGS as BLUP PGS.

#### Evaluation of prediction accuracy

For each phenotype, we used a 5-fold cross-validation scheme to obtain out-of-sample prediction accuracy estimates. The prediction was evaluated by 1) adjusted variance explained in the liability scale. We used population prevalences specified in ST3. to convert the variance explained in a linear regression to the liability scale^51^. The adjusted R2 was defined as the variance explained by the full model after accounting for the variance explained by the base model as R2_adjusted = (R2_full - R2_cov) / (1 - R2_cov). 2) Odds ratio (OR) of the 5th quintile to the middle quintile in the case of risk scores and 3) OR of the 10th decile to the 1st decile in the case of polygenic scores only. All OR were calculated from a logistic regression model based on the PGS percentiles, sex, birth year and first 20PCs. 4) Pearson’s correlation estimates.

## Data Availability

All relevant iPSYCH and Danish ANGI data are available from the authors after approval by the iPSYCH Data Access Committee and can only be accessed on the secured Danish server as the data are protected by Danish legislation. More information about getting access to the Danish data can be obtained at http://ipsych.au.dk/about-ipsych/. We utilized the list of GWAS Catalog (https://www.ebi.ac.uk/gwas/) summary statistics downloaded on 09/09/2020, GWAS summary statistics from the PGC (https://www.med.unc.edu/pgc/download-results/) and GWAS Atlas UKB2 data freeze v20191115 (https://atlas.ctglab.nl/).

## Acknowledgements

C.A., B.J.V. and F.P. were supported by the Danish National Research Foundation (Niels Bohr Professorship to Prof. John McGrath), the Lundbeck Foundation Initiative for Integrative Psychiatric Research, iPSYCH (R102-A9118, R155-2014-1724 and R248-2017-2003), and a Lundbeck Foundation Fellowship (R335-2019-2339). C.A. was also supported by a Willam Demant Fonden travel grant. High-performance computer capacity for handling and statistical analysis of iPSYCH data on the GenomeDK HPC facility was provided by the Center for Genomics and Personalized Medicine and the Centre for Integrative Sequencing, iSEQ, Aarhus University, Denmark (grant to A.D.B.). IB was also supported by the Swedish Brain Foundation and Fredrik och Ingrid Thurings Stiftelse. AN data are from the Anorexia Nervosa Genetics Initiative, an initiative of the Klarman Family Foundation, and extendented with support from the Lundbeck foundation (R276-2017-4581).

## Code availability

All code used in this project is available in GitHub (https://github.com/ClaraAlbi/paper_multiPGS). We used the following tools: BOLT-LMM (https://data.broadinstitute.org/alkesgroup/bolt-lmm), XGBoost (https://xgboost.readthedocs.io/en/latest/), SMTpred (https://github.com/uqrmaie1/smtpred), bigsnpr/LDpred2 (https://privefl.github.io/bigsnpr/articles/LDpred2.html).

