## Supplementary material for "Multi-PGS enhances polygenic prediction: weighting 937 polygenic scores"


**Supplementary text** 2

PGS library construction 2

1- GWAS summary statistics download 2

2- Parse and format the GWAS summary statistics 2

3- Quality control 4

4- Run LDpred2-auto 4

References 6

**Supplementary tables** 7

ST2. Meta-data and results PGS library. 7

ST3. Summary of main iPSYCH outcomes and compared PGS. 7

ST4. List of all phenotypes and ICD10 codes. 8

ST5. Summary of other outcomes and compared PGS. 8

**Supplementary figures** 8

SF5. AUC iPSYCH 8

SF6. ADHD multiPGS lasso weights 10

SF7. AFF multiPGS lasso weights 11

SF8. AN multiPGS lasso weights 12

SF9. ASD multiPGS lasso weights 13

SF10. BD multiPGS lasso weights 15

SF11. SCZ multiPGS lasso weights 16

SF12. Properties of the GWAS summary statistics for top 5 PGS 18

SF13. ICD10 multiPGS prediction 19

### Supplementary text

#### PGS library construction

##### 1- GWAS summary statistics download

The following resources were used to obtain the GWAS summary statistics for generating PRSs:

| Source | Number of files |
|---|---|
| GWAS Catalog[1] data freeze 2020-09-09 | 1376 |
| GWAS ATLAS[2] data freeze v20191115 | 4756 |
| PGC (exclude iPSYCH samples) | 12 |
| Other public sources | 62 |

**Table S1. Overview of sources for the initial list of GWAS summary statistics.**

For the first two data resources, the following filtering was used:

- Total sample > 10,000.
- Number of SNPs in the file > 250,000.
- Training sample of European ancestry. Variables "BROAD ANCESTRAL CATEGORY" for the GWAS Catalog and Population "EUR" or "UKB2 (EUR)" for the GWAS ATLAS.
- For GWAS of the same phenotype, the latest addition to the catalog was selected.
- For GWAS ATLAS, only data from 2014 forward was included.

For the specific PGC GWAS summary statistics where iPSYCH was used in the discovery dataset, we used in-house GWAS results where these samples were excluded from the calculation. **The GWAS were specifically selected to be based on European ancestry individuals, to not be overly redundant and to not contain iPSYCH samples (manually checked).**

For more details, check the script (github script 1-prepare-sumstats.R) that defines the list of GWAS summary statistics from GWAS Catalog and GWAS ATLAS.

**Resulting # files: 1,377**

For more details, check the script (github script 1-prepare-sumstats.R) that downloaded, read and stored the header and metadata for each GWAS summary statistic file.

##### 2- Parse and format the GWAS summary statistics

We constructed an internal library of GWAS summary statistics header possibilities / column names and filtered out the files with missing essential categories (Figure S1). The essential categories considered were:

**chr**: chromosome
**rsid**: rsID number or **pos**: base pair position under hg19
**a1**: effect allele
**a0**: no-effect allele
**beta**: linear / logistic regression effect size
**or**: odds ratio
**beta_se**: standard error of effect size
**p**: p-value

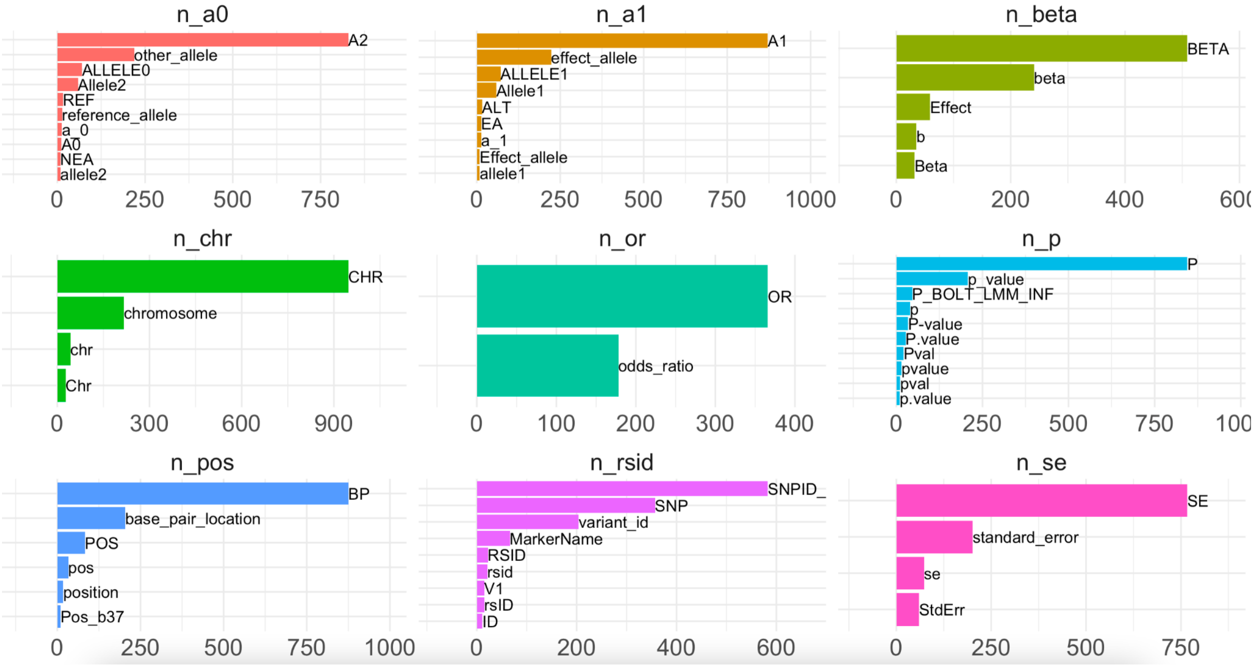

**SF1. Most frequently used column names for the essential categories.** Filtered for n>8.

Processing/filters applied:

- Empty beta / or columns or > 30% of the file length is empty ▶️ full file discarded.
- rsid column format can't be processed (different string processing tried) ▶️ full file discarded.
- Create an effective sample size (required by LDpred2 [3], Neff = 4/(1/Ncases + 1/Ncontrol)) column from reading the file (if available), otherwise use metadata.
- Check if the standard error column is from the beta, the log(beta) or the odds ratio and transform if required.
- Check median chi2 statistic (beta/beta_se)^2 for formatted file. If very large ▶️ full file discarded.
- Calculate LDSC regression[4] SNP-h2 (prior h2 parameter for LDpred2-auto) and intercept estimates.

**Resulting # files: 1,005**

For more details, check the script (github script 2-parser.R) that parses and re-formats each GWAS summary statistic file.

#### 3- Quality control

As recommended in the LDpred2 paper[5], filtering SNPs with a large discrepancy in standard deviations between the genotyped/imputed data and the GWAS summary statistics increases the prediction accuracy of PRSs.

Processing/filters applied:

- Filter SNPs to HapMap3 set of variants.
- Filter SNPs to match LD reference set (https://figshare.com/articles/dataset/European_LD_reference_with_blocks_/19213299).
- SNP QC step described in Privé *et al.* 2020[6]
- Make a quality control plot (example on SF2).
- Check the number of resulting SNPs. If < 200,000 ▶️ full file discarded.

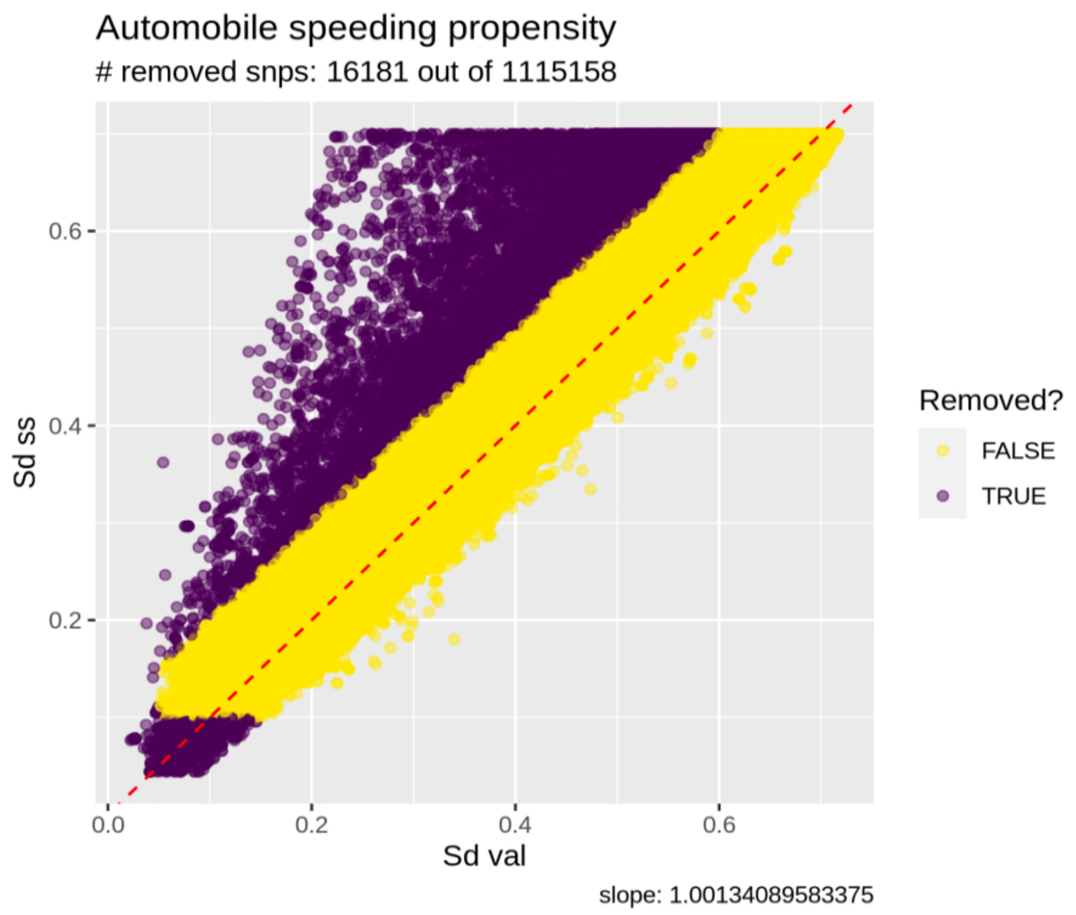


**SF2. Example QC plot.** The trait shown is for PMID 30643258[7] GWAS summary statistics (Automobile speeding propensity).

**Resulting # files: 952**

For more details, check the script (github script 3-qc.R) that controls and restricts the number of SNPs in each GWAS summary statistic file.

#### 4- Run LDpred2-auto

Polygenic risk scores were derived using LDpred2-auto, a method within the LDpred2 framework[3] that does not require a validation dataset to fit the hyperparameters (SNP-h2; SNP-based heritability estimate and p; proportion of causal SNPs), but these are fitted as part of the Gibbs sampler instead. We used the provided European-ancestry independent LD blocks as reference panel[6]. For each GWAS summary statistics file, LDpred2-auto was run with 30 Gibbs sampler chains, 800 burn-in iterations and 400 iterations. The SNP-h2 initial value was set to the LD score regression estimate[8] from the GWAS summary statistics after QC. Each of the chains was initialized with a different value for the proportion of causal variants: in the range [1e-4, 0.9], equally spaced on a log scale. Chains were filtered according to the recommendation in the LDpred2 tutorial, and effect sizes of chains kept were averaged (example convergence plot on Figure S3).

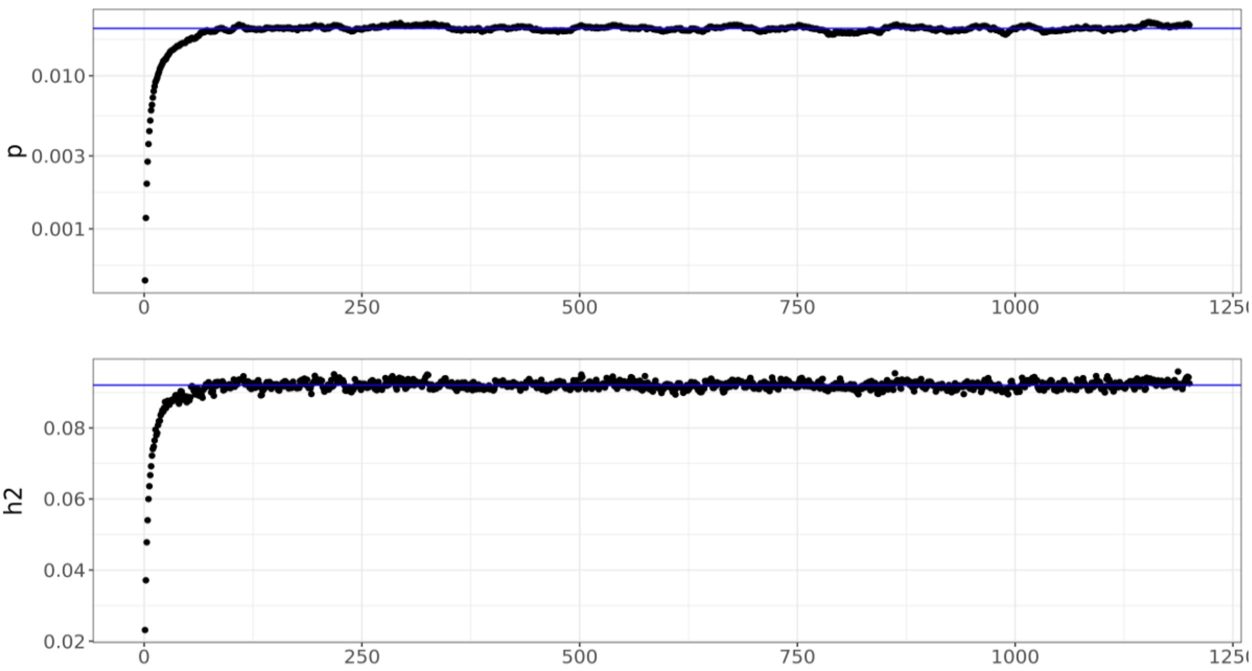


**SF3. Example convergence plot for LDpred2-auto.**

Processing/filters applied:

- The LDpred2-auto algorithm did not converge ▶️ full file discarded.
- Estimated h2 from LDpred2-auto > 1 ▶️ full file discarded.

**Resulting # files: 937**

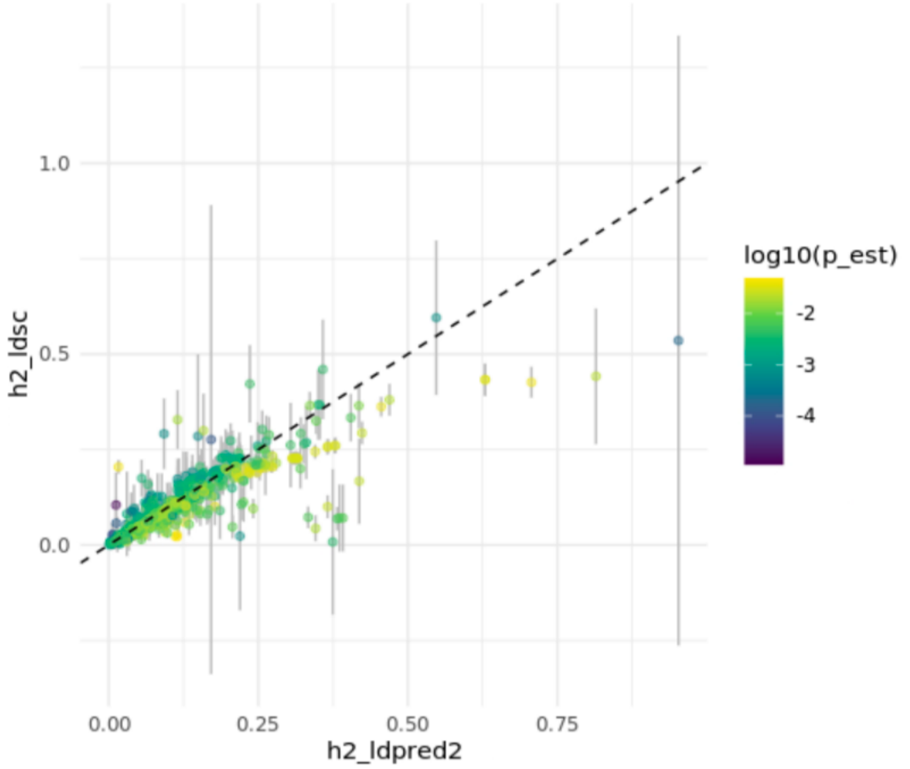


**Figure S4. Results for the 937 LDpred2-auto polygenic scores in our PGS library.**

For more details, check the script (github script 4-run-ldpred2.R) for code on running LDpred2-auto.

### Supplementary tables

#### ST2. Meta-data and results PGS library.

Information on the 937 GWAS summary statistics with LDpred2-auto PGS that were the input to the multiPGS models. GWAS_trait: reported outcome of GWAS, id: unique identifier of GWAS, GWAS_pubmedID: PubMed ID of GWAS, source: file downloaded from this resource, M_or: original number of variants in GWAS summary statistics file, M_m: number of variants that matches the iPSYCH HM3 subset, M_ldpred: number of variants that matches the LDpred2 provided LD reference for European ancestry and passes QC (described in Supplementary Text), k: number of Gibbs sampler chains that passes the LDpred2 recommended filter (out of 30), h2_est: LDpred2-auto estimated SNP-heritability parameter, p_est: LDpred2-auto estimated proportion of causal variants parameter.
https://github.com/ClaraAlbi/paper_multiPGS/blob/main/supplementary_data/supplementary_data_2.csv

#### ST3. Summary of main iPSYCH outcomes and compared PGS.

Information on the 6 psychiatric disorders: attention-deficit/hyperactivity disorder (ADHD), affective disorder (AFF), anorexia nervosa (AN), autism spectrum disorder (ASD), bipolar disorder (BD) and schizophrenia (SCZ) in the main analysis. Tag: outcome label, p: ICD10 category, cv: 5-fold cross-validation subset, prev_pop: population prevalence used for liability-scale transformation, prev_gwas: individual-level data case-control ratio used for liability-scale transformation, n_cases_train: number of cases in training subset, n_control_train, number of controls in training subset, n_cases_test: number of cases in testing subset, n_control_test: number of controls in testing subset, comp: label for the GWAS summary statistics used to compute the PGS to compare against, GWAS_trait, compared outcome, GWAS_pubmedID: PubMed ID of GWAS, id: unique identifier of GWAS for

meta-data PGS library, n_cases_GWAS: number of cases in GWAS, n_control_GWAS: number of controls in GWAS.
https://github.com/ClaraAlbi/paper_multiPGS/blob/main/supplementary_data/supplementary_data_3.csv

#### ST4. List of all phenotypes and ICD10 codes.

First cross-validation subset information for all used phenotypes and ICD10 codes in the analyses, including information on the sample overlap with the iPSYCH cohort and the main diagnosis. p: ICD10 category, diagnosis: description of the ICD10 category, cases: number of cases in first cross-validation subset, scz, adhd, asd, aff, bip, cohort: proportion of cases that overlap with the different groups.
https://github.com/ClaraAlbi/paper_multiPGS/blob/main/supplementary_data/supplementary_data_4.csv

#### ST5. Summary of other outcomes and compared PGS.

Information on the 15 traits selected for Figure 5. For ADHD/ASD, ASD was coded as 0 (controls) and ADHD was coded as 1 (cases). For BD/MDD, MDD was coded as 0 (controls) and BD was coded as 1 (cases). Tag: outcome label, p: ICD10 category, cv: 5-fold cross-validation subset, prev_pop: population prevalence used for liability-scale transformation, prev_gwas: individual-level data case-control ratio used for liability-scale transformation, n_cases_train: number of cases in training subset, n_control_train, number of controls in training subset, n_cases_test: number of cases in testing subset, n_control_test: number of controls in testing subset, comp: label for the GWAS summary statistics used to compute the PGS to compare against, GWAS_trait, compared outcome, GWAS_pubmedID: PubMed ID of GWAS, id: unique identifier of GWAS for meta-data PGS library, n_cases_GWAS: number of cases in GWAS, n_control_GWAS: number of controls in GWAS.
https://github.com/ClaraAlbi/paper_multiPGS/blob/main/supplementary_data/supplementary_data_5.csv

### Supplementary figures

#### SF5. AUC iPSYCH

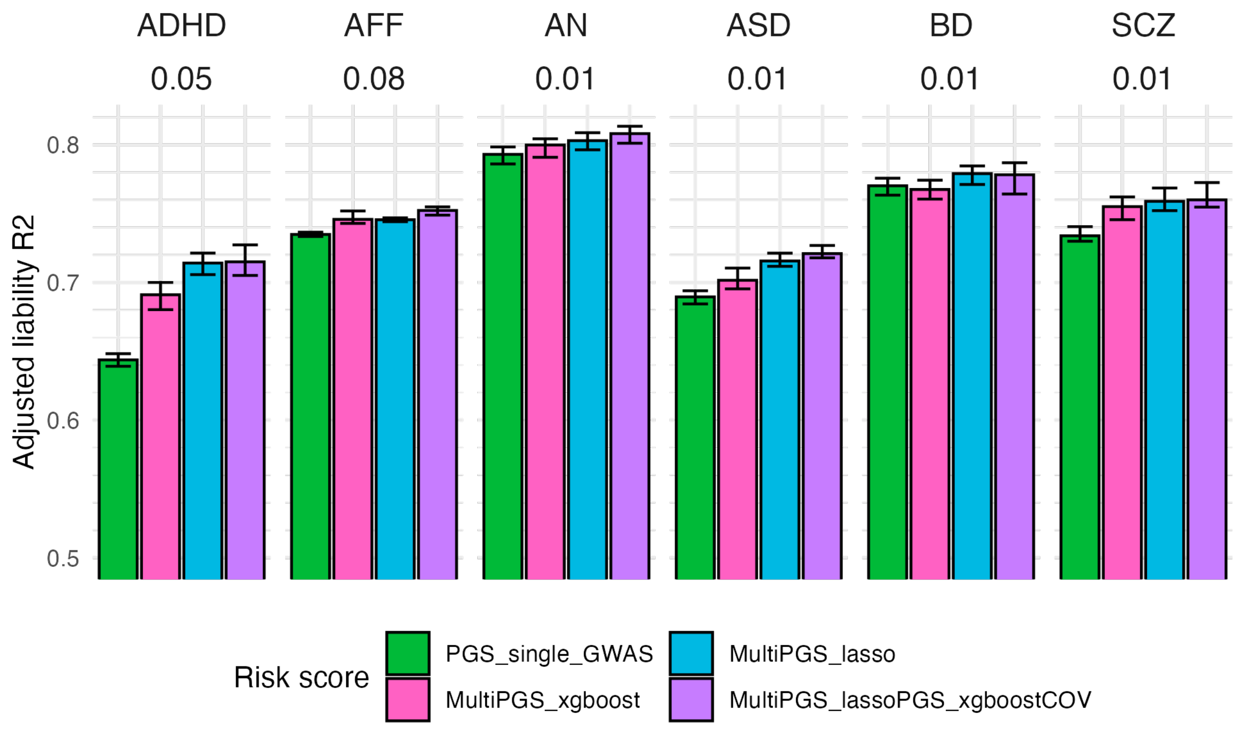


**SF5. Performance of the different risk score models including covariates.** Comparison between the per-disorder attention-deficit/hyperactivity disorder (ADHD), affective disorder (AFF), anorexia nervosa (AN), autism spectrum disorder (ASD), bipolar disorder (BD) and schizophrenia (SCZ) single GWAS PGS (specific details on ST3) and the multi-PRS models trained with 937 PGS in terms of AUC. All models included sex, age and first 20 PCs for training the different PGS weights and calculating the risk score on the test set in a 5-fold cross-validation scheme. Confidence intervals were calculated from 10,000 bootstrap samples of the mean adjusted AUC.

### SF6. ADHD multiPGS lasso weights

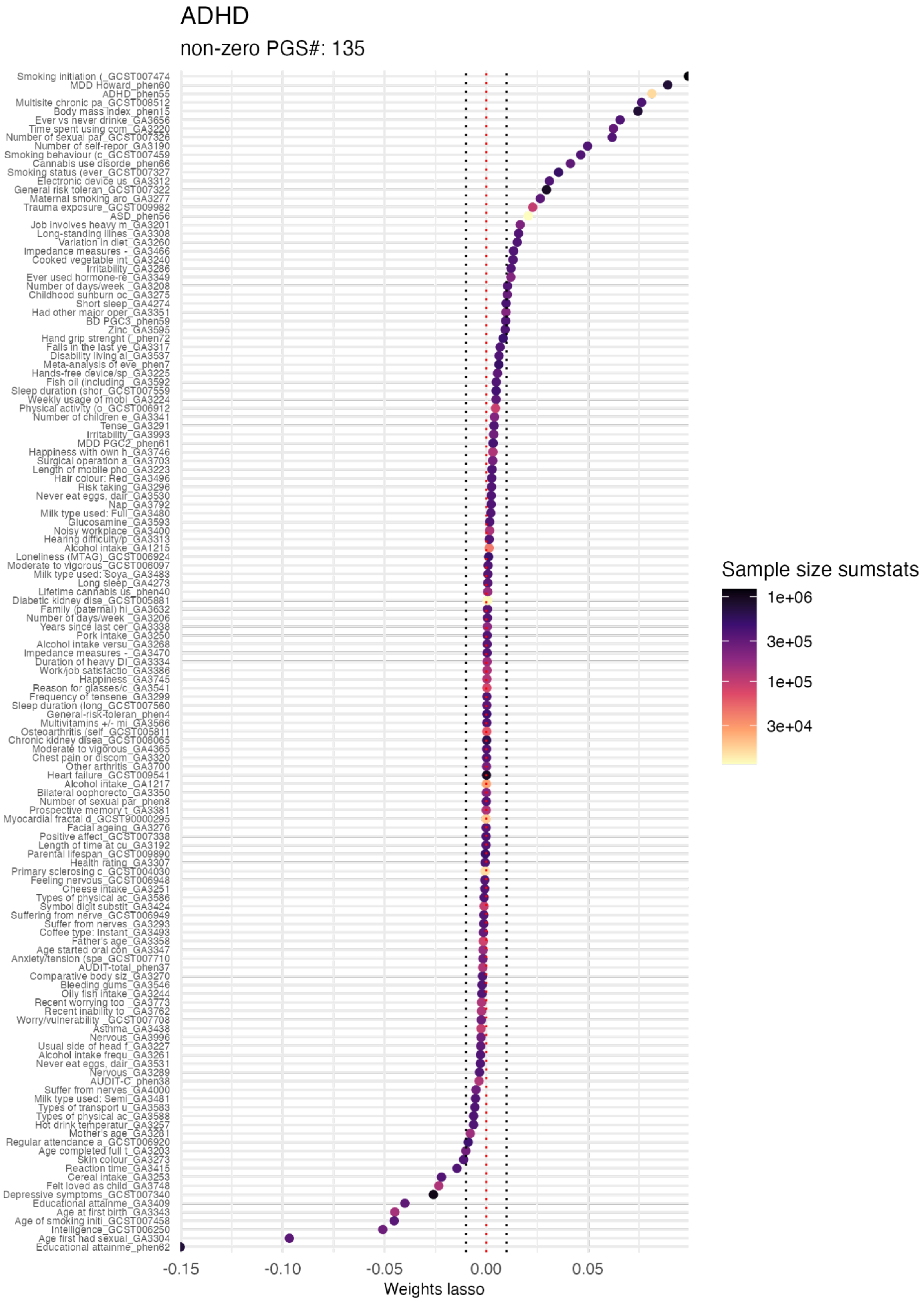


**SF6. ADHD multiPGS lasso weights.** Mean lasso weight (x-axis) for the 5 cross-validation subsets of the lasso multiPGS. Each PGS with a non-zero lasso weight is represented by the y-axis with GWAS outcome name and the unique GWAS identifier (ST2). The color indicates the sample size of the external GWAS summary statistics for each PGS. The number of variables in the y-axis is available in the subtitle.

#### SF7. AFF multiPGS lasso weights

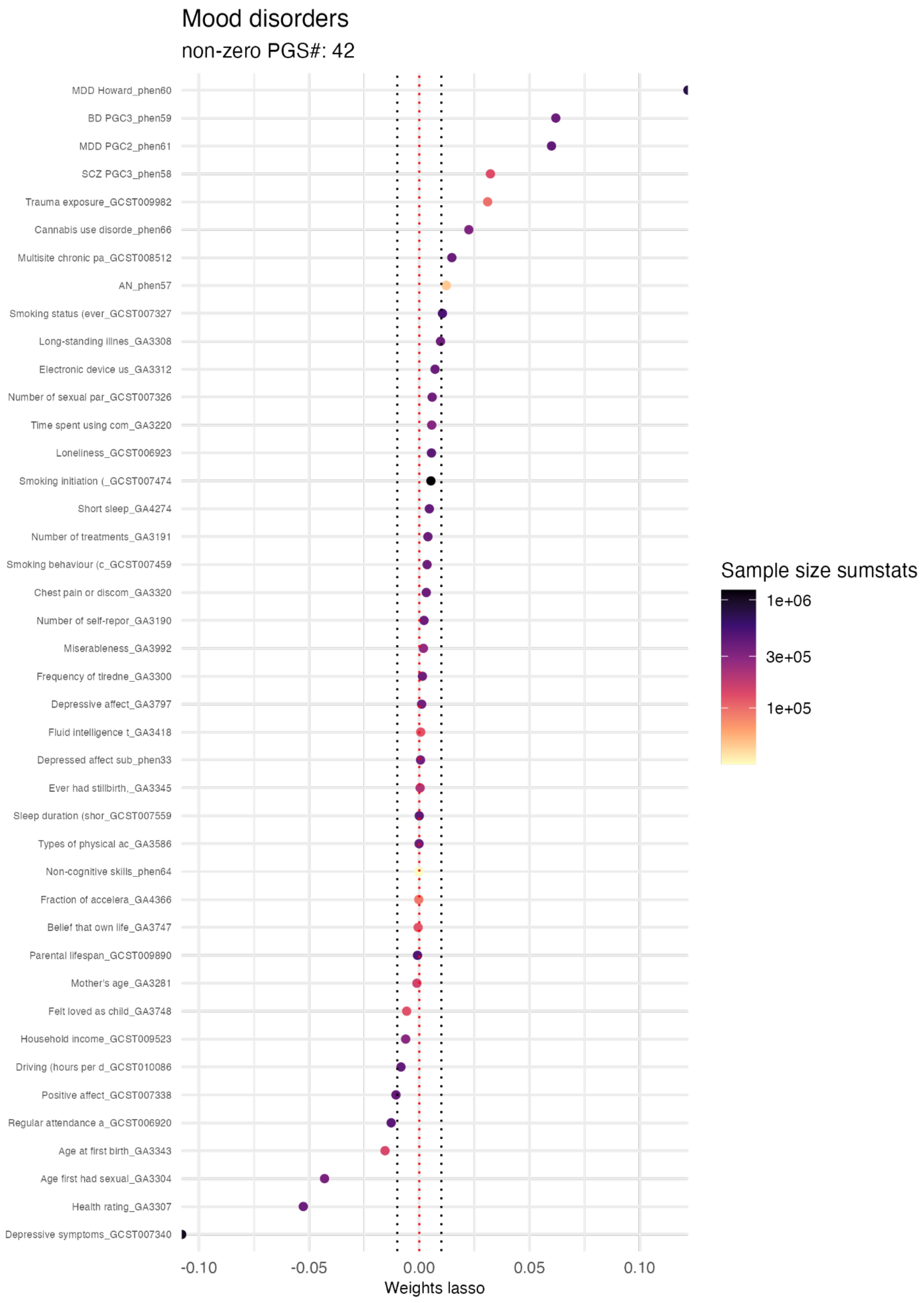


**SF7. AFF multiPGS lasso weights.** Mean lasso weight (x-axis) for the 5 cross-validation subsets of the lasso multiPGS. Each PGS with a non-zero lasso weight is represented by the y-axis with GWAS outcome name and the unique GWAS identifier (ST2). The color indicates the sample size of the

external GWAS summary statistics for each PGS. The number of variables in the y-axis is available in the subtitle.

#### SF8. AN multiPGS lasso weights

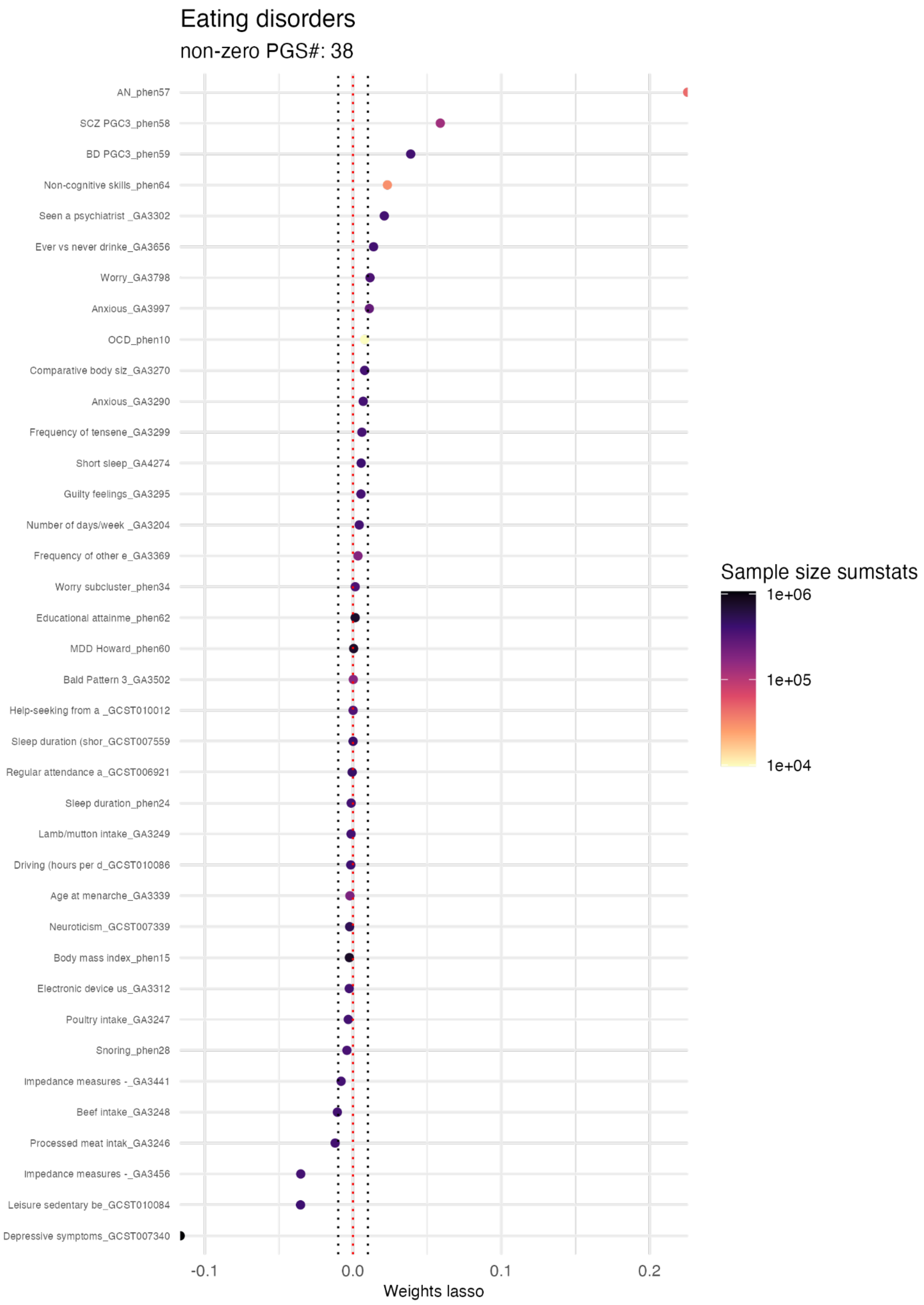


**SF8. AN multiPGS lasso weights.** Mean lasso weight (x-axis) for the 5 cross-validation subsets of the lasso multiPGS. Each PGS with a non-zero lasso weight is represented by the y-axis with GWAS

outcome name and the unique GWAS identifier (ST2). The color indicates the sample size of the external GWAS summary statistics for each PGS. The number of variables in the y-axis is available in the subtitle.

#### SF9. ASD multiPGS lasso weights

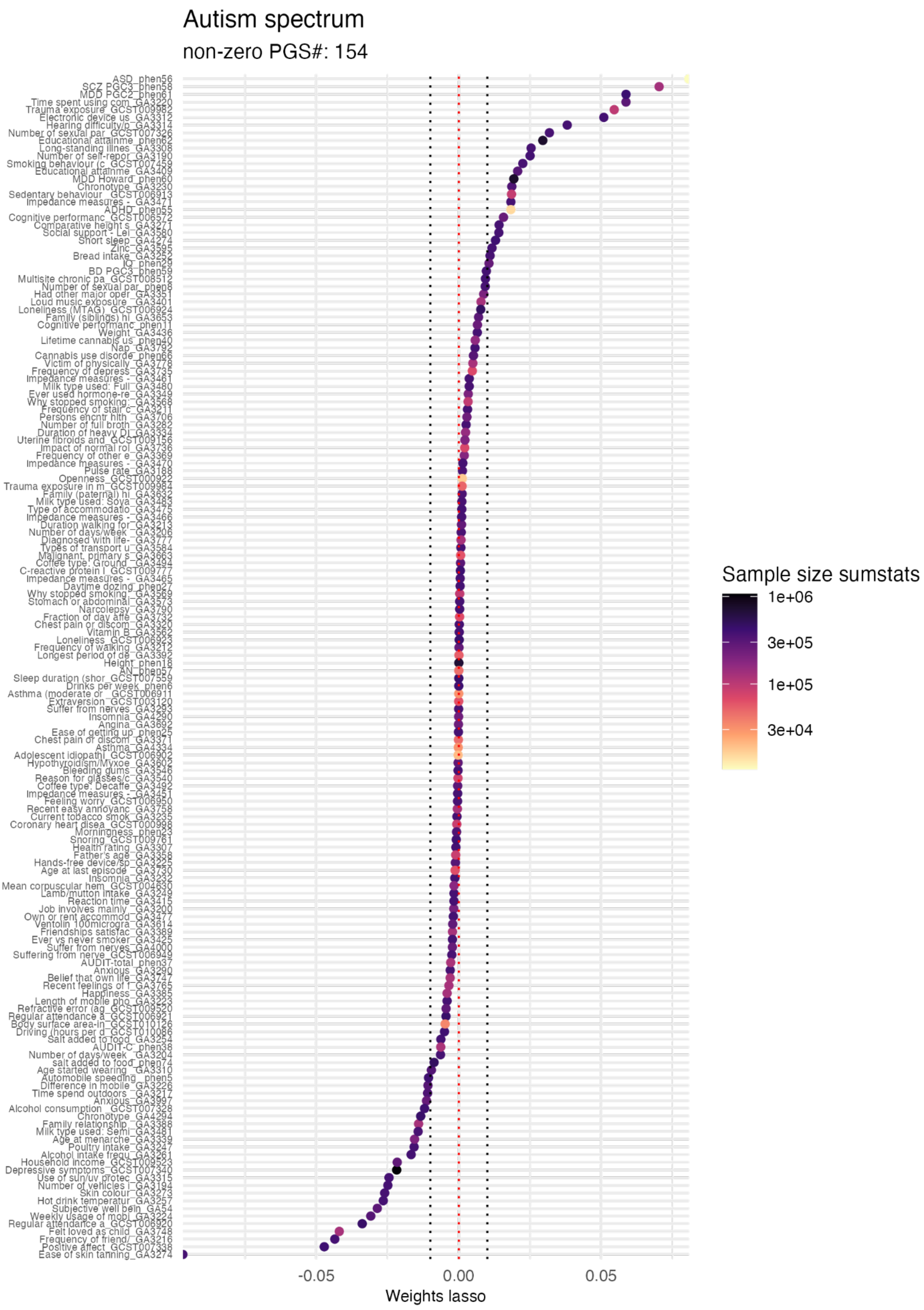


**SF9. ASD multiPGS lasso weights.** Mean lasso weight (x-axis) for the 5 cross-validation subsets of the lasso multiPGS. Each PGS with a non-zero lasso weight is represented by the y-axis with GWAS outcome name and the unique GWAS identifier (ST2). The color indicates the sample size of the external GWAS summary statistics for each PGS. The number of variables in the y-axis is available in the subtitle.

#### SF10. BD multiPGS lasso weights

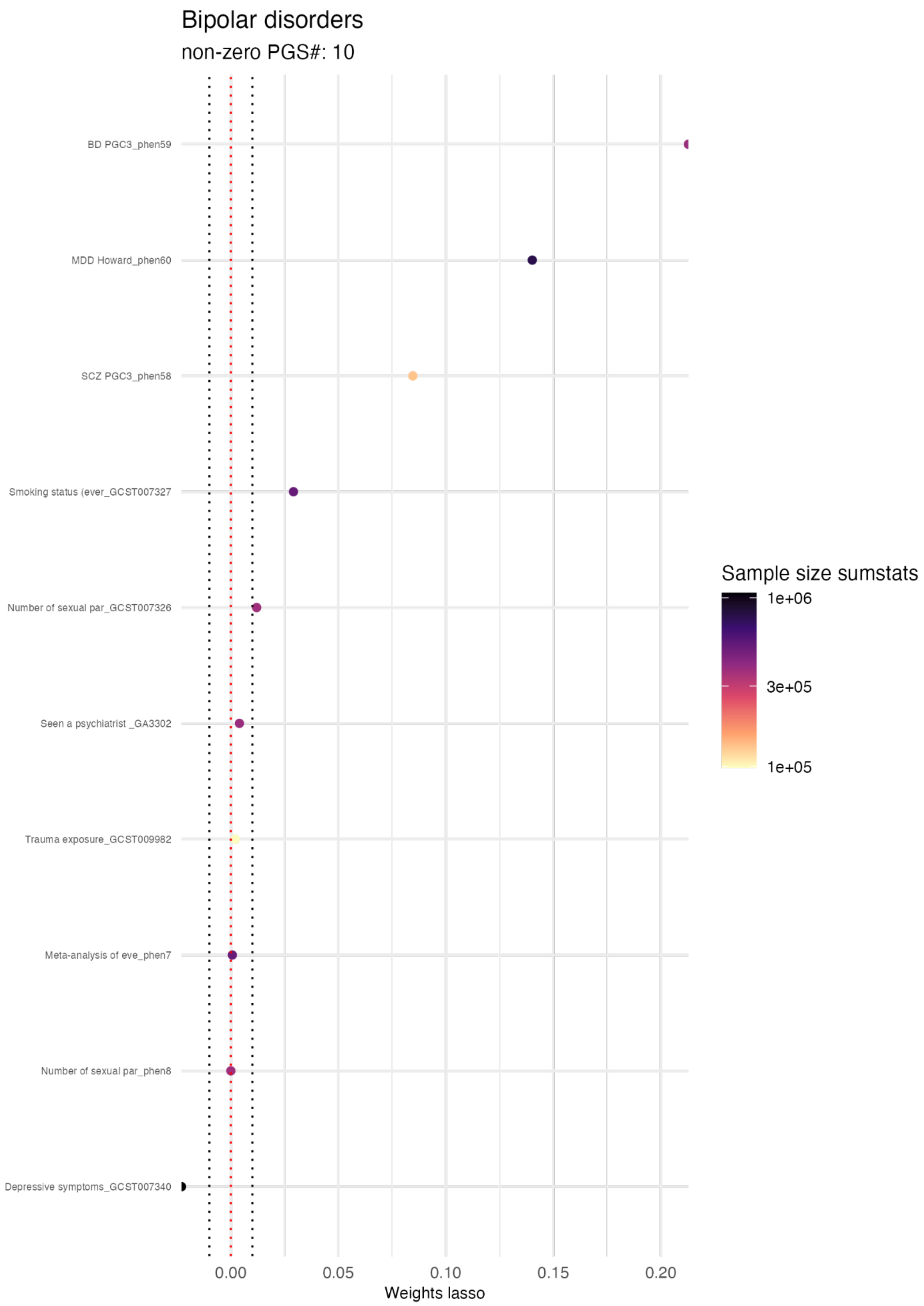


**SF10. BD multiPGS lasso weights.** Mean lasso weight (x-axis) for the 5 cross-validation subsets of the lasso multiPGS. Each PGS with a non-zero lasso weight is represented by the y-axis with GWAS outcome name and the unique GWAS identifier (ST2). The color indicates the sample size of the external GWAS summary statistics for each PGS. The number of variables in the y-axis is available in the subtitle.

#### SF11. SCZ multiPGS lasso weights

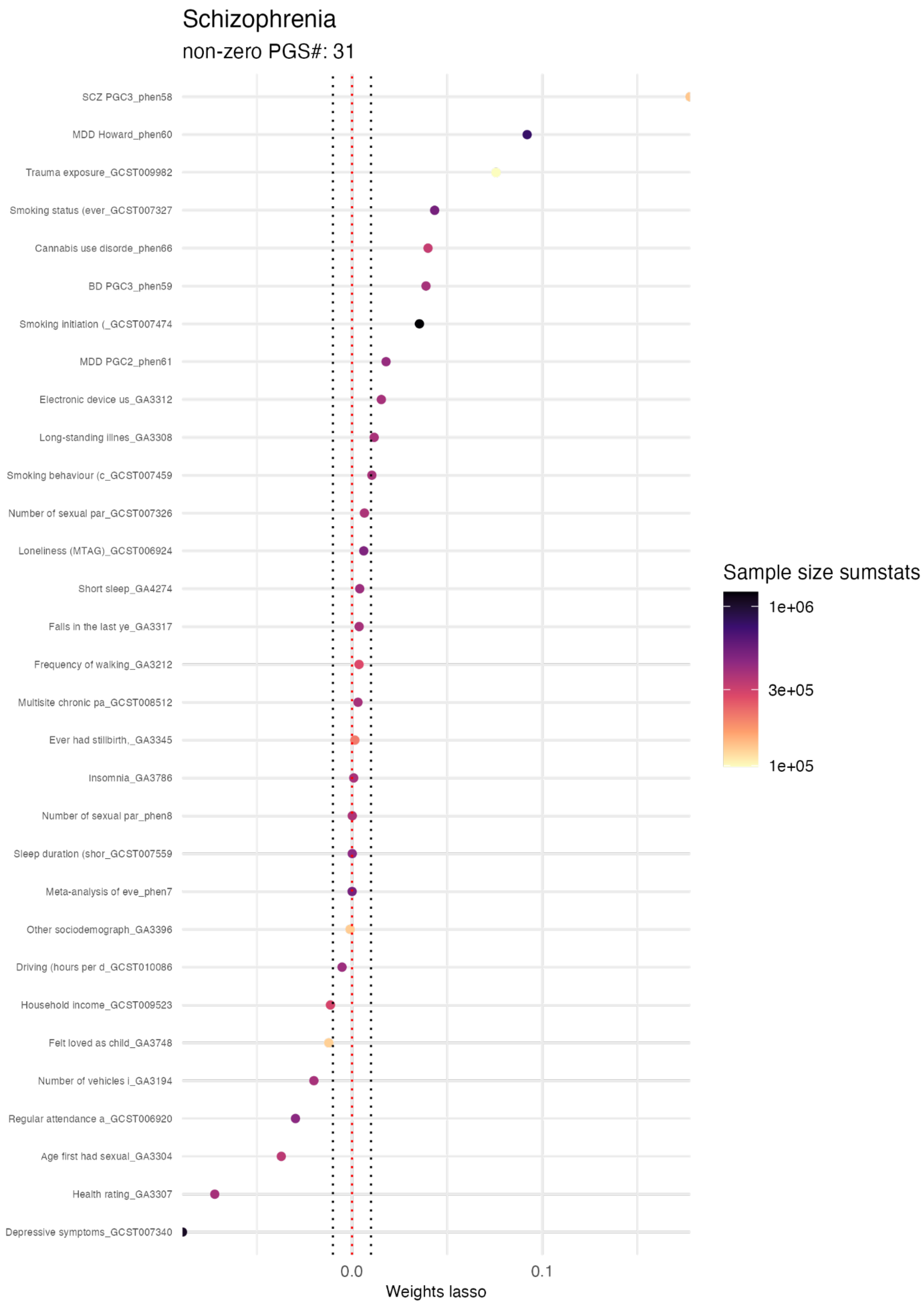


**SF11. SCZ multiPGS lasso weights.** Mean lasso weight (x-axis) for the 5 cross-validation subsets of the lasso multiPGS. Each PGS with a non-zero lasso weight is represented by the y-axis with

GWAS outcome name and the unique GWAS identifier (ST2). The color indicates the sample size of the external GWAS summary statistics for each PGS. The number of variables in the y-axis is available in the subtitle.

### SF12. Properties of the GWAS summary statistics for top 5 PGS

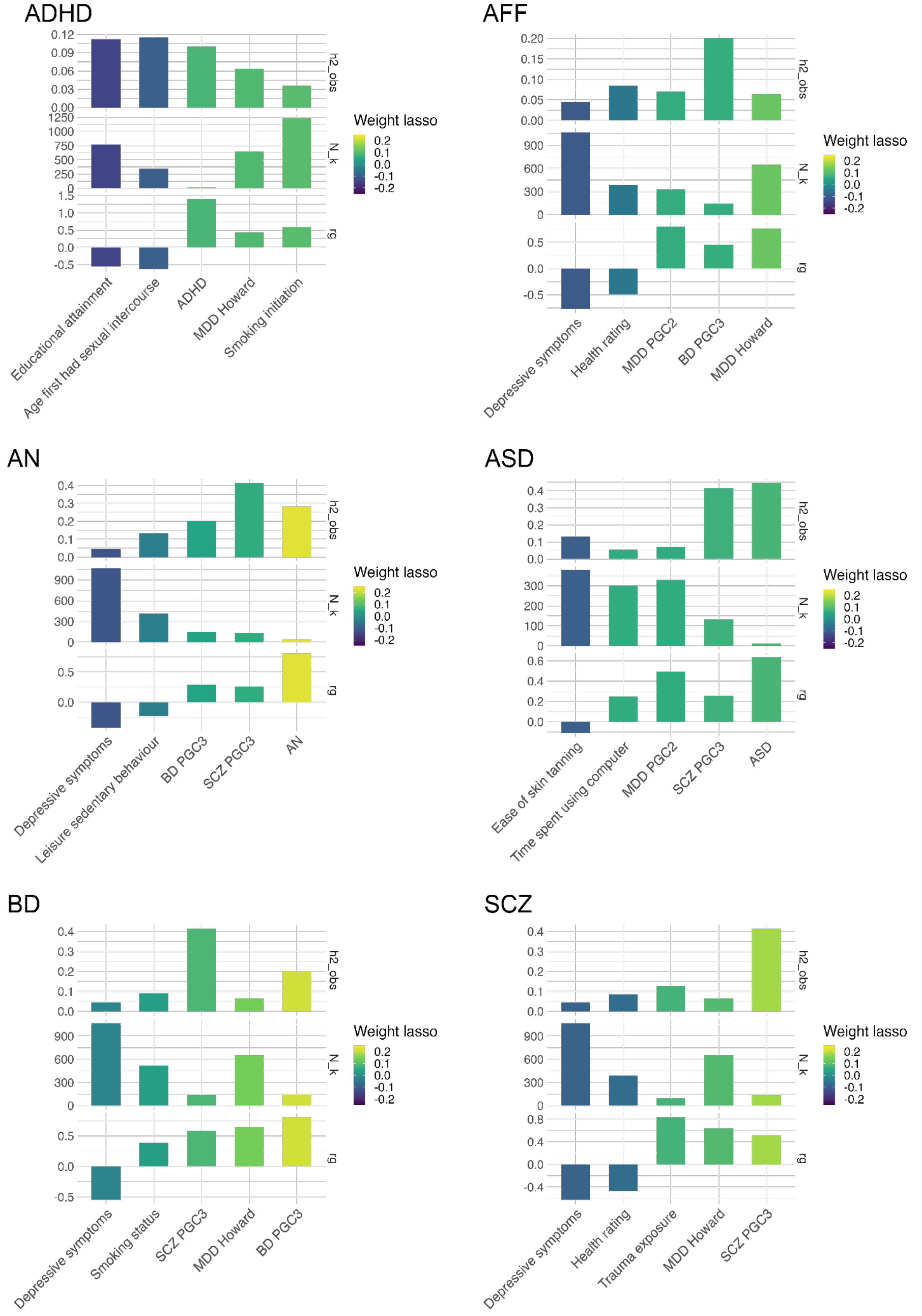


**SF12. Properties of the GWAS summary statistics for top 5 PGS.** Top 5 PGS based on the lasso weight (x-axis) for attention-deficit/hyperactivity disorder (ADHD), affective disorder (AFF), anorexia

nervosa (AN), autism spectrum disorder (ASD), bipolar disorder (BD) and schizophrenia (SCZ), with SNP heritability in the observed scale (h2_obs), sample size in thousands (N_k) and genetic correlation with the predicted trait (rg) (from top to bottom in each panel).

#### SF13. ICD10 multiPGS prediction

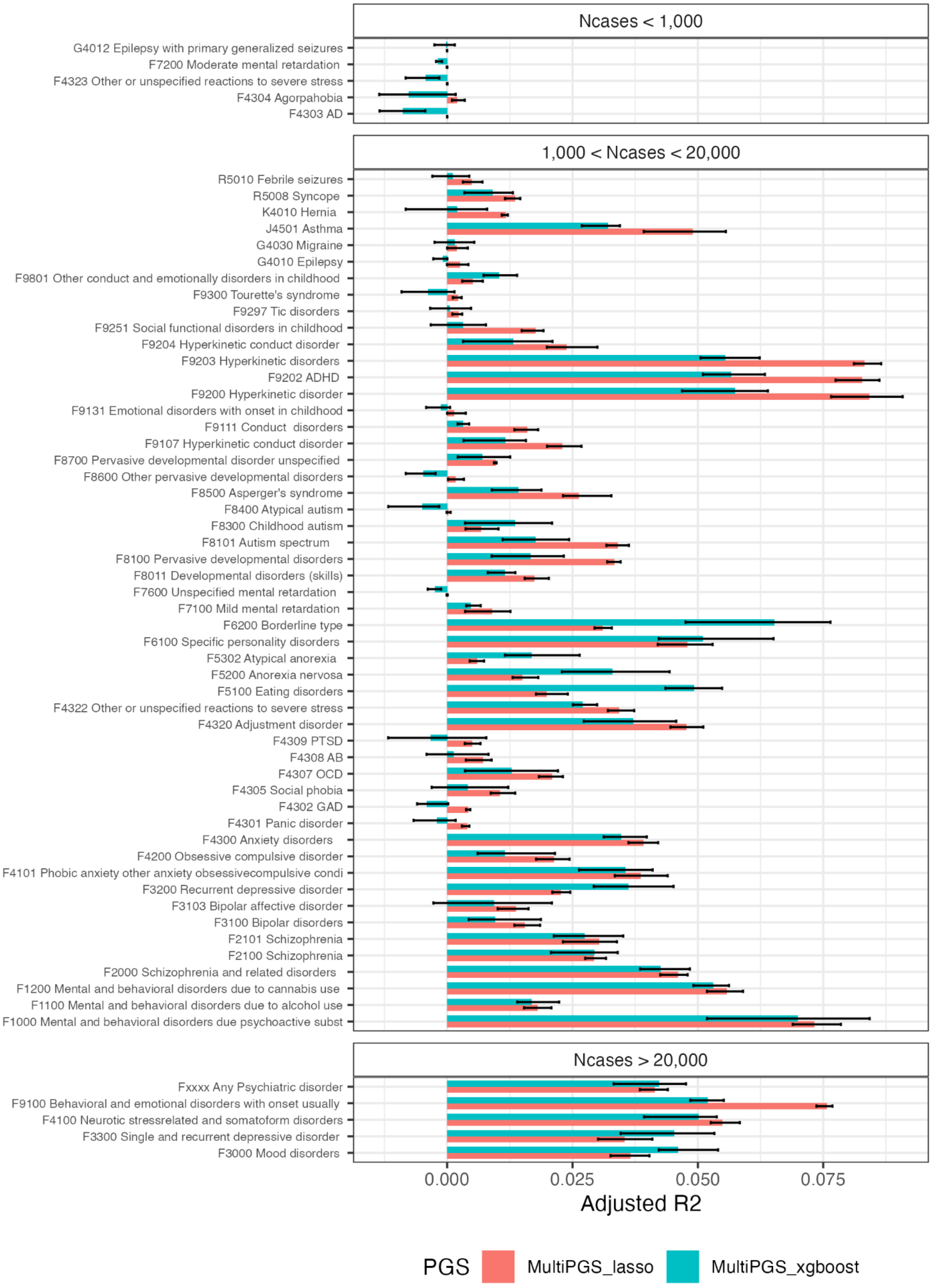


**SF13. Performance of the multi-PGS on register-based outcomes.** Comparison between the per-disorder multi-PRS models trained with 937 PGS adjusted R2. All models included sex, age and first 20 PCs for training the different PGS weights and calculating the risk score on the test set in a 5-fold

cross-validation scheme. Confidence intervals were calculated from 10,000 bootstrap samples of the mean adjusted R2, where the adjusted R2 was the variance explained by the full model after accounting for the variance explained by a logistic regression covariates-only model as R2_adjusted = (R2_full - R2_cov) / (1 - R2_cov).
